# Unlocking the Potential of Genetic Research in Pulmonary Arterial Hypertension: Insights from Clinicians, Researchers and Study Team

**DOI:** 10.1101/2023.10.27.23297700

**Authors:** Emilia M Swietlik, Michaela Fay, Nicholas W Morrell

## Abstract

**Background:** While genetics has been incorporated into various subspecialties of medicine for some time, the significance of genetic research and testing in understanding the fundamental mechanisms of Pulmonary Arterial Hypertension (PAH) and formulating effective treatment approaches has only recently come to the forefront. We aimed to explore how attitudes toward genetic research among clinical and research teams impacted the engagement in genetic research and the integration of genetic insights into clinical practice.

**Methods:** Participants were selected from the National Institute for Health Research BioResource Rare Diseases study (NBR) and the Cohort study of idiopathic and heritable PAH (the PAH Cohort), representing a range of roles, ages, genders, and mutation statuses. We conducted a total of 53 semi-structured interviews and focus groups involving a total of 63 patients, clinicians, and researchers from nine UK PH centres. Following the principles of Grounded Theory, interview transcripts were thematically coded by two authors using MAXQDA (2022) software. In this paper, we focus on the researchers’, clinicians’ and study team’s perspectives.

**Results:** From the interview data, several key themes emerged, ranging from study design, recruitment and consent procedures to the return of individual genetic results. Additionally, participants reflected on both the successes of these studies and the future directions of genetic research. The analysis highlighted the critical importance of fostering collaborative networks firmly rooted in existing clinical and research infrastructure in rare disease study setups. Furthermore, the significance of trust-building, personalised communication, and transparency among stakeholders was underscored. The study offered valuable insights into the motivating and hindering factors to participant recruitment and consent procedures. Lastly, the findings gathered from processes surrounding the return of individual genetic results, genetic counselling, and the recruitment of relatives provided invaluable lessons regarding the integration of genetics into clinical practice.

**Conclusions:** This in-depth analysis yields a crucial understanding of attitudes to genetic research among various stakeholders and sheds light on the complexities of genetic research and the evidence-practice gap.

## Introduction

While genetics has played a significant role in various subspecialties of medicine, its emergence as a vital tool in understanding Pulmonary Arterial Hypertension (PAH) and developing effective treatments is relatively recent. Two substantial studies, the National Institute for Health Research BioResource Rare Diseases study (NBR) and the Cohort study of idiopathic and heritable PAH (the PAH Cohort), have been pivotal in this endeavour. These studies employed whole genome sequencing (WGS) and deep phenotyping, encompassing both adult and paediatric incidents and prevalent cases, along with their relatives. This collaborative effort has resulted in the largest and most comprehensive cohort to date for individuals with I/HPAH, PVOD, and PCH and their relatives. While the NBR study has already returned pertinent individual genetic results to patients who consented, the PAH Cohort study has not. The studies led to the discovery of new genetic variants associated with the disease^1–4^ and provided insights into its pathology using omic technologies^5–8^, but their successes extend far beyond their scientific output. As the NBR study concluded and the PAH Cohort study reached its 10-year milestone, we set out to ask clinicians, researchers, and the study team about their experience of these studies and the subsequent steps for implementing research findings into clinical practice.

### Aims and Objectives

The RAPID-PAH study aimed to explore attitudes toward genetic research, assess clinicians’ abilities and confidence in explaining genetic research findings and examine their influence on the pace, success, and patient and relative participation in genetic studies. Ultimately, this research seeks to understand how these factors impact the integration of genetic insights into clinical practice.

### Design and Methods

A purposive sample of participants from the NBR and PAH Cohort study was selected with a view to include/represent diverse age groups, genders, and individuals with and without PAH risk gene mutations. The recruitment process covered all PH centres in the UK and took four distinct approaches (Table 1), including contacting individuals through local clinical or research teams, referrals from current participants, online recruitment via email, social media platforms, and the Pulmonary Hypertension Association (PHA) webpage. Active recruitment continued until the target number of participants had been attained (Figure 1). Clinical, research and study teams were contacted directly.

**Figure 1.**
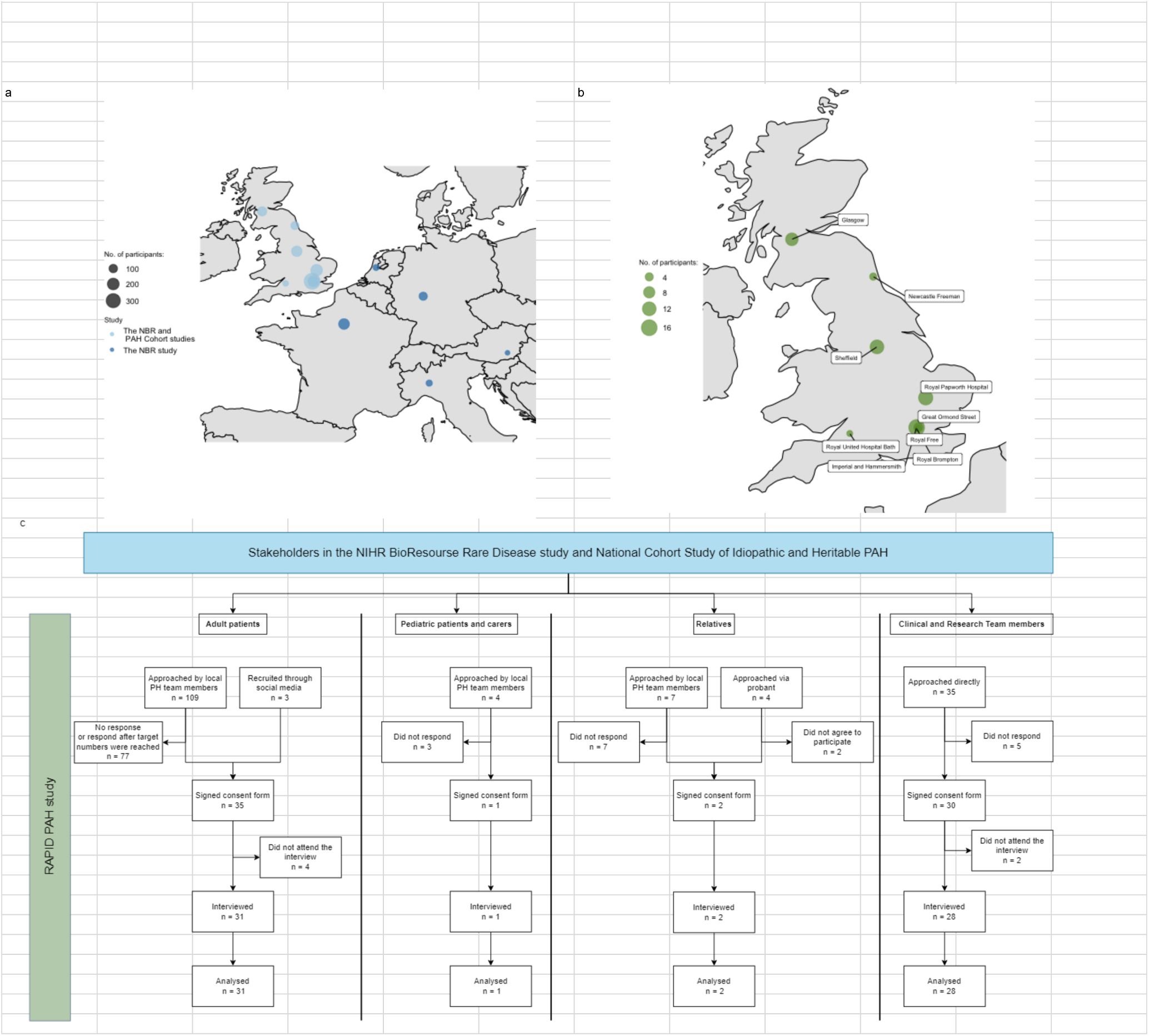
Depiction of the geographical distribution of participants in (a) the NIHR BioResource Rare Disease study and the National Cohort Study of Idiopathic and Heritable Pulmonary Arterial Hypertension (PAH), (b) in the RAPID-PAH study. The size of the plotting character represents the participant count, (c) Consort diagram of RAPID PAH.

**Table 1.** Demographic characteristics of participants.

| **Clinical and Research Teams N=28** |  |
| --- | --- |
| **Role:** |  |
| Clinical Trial Coordinator | 3 (11%) |
| Doctor | 15 (54%) |
| Research Assistant | 1 (4%) |
| Research Nurse | 4 (14%) |
| Researcher | 3 (11%) |
| Specialist Nurse | 2 (7%) |
| **Centre:** |  |
| Great Ormond Street | 4 (14%) |
| Imperial and Hammersmith | 6 (21%) |
| Newcastle Freeman | 2 (7%) |
| NHS Greater Glasgow and Clyde | 3 (11%) |
| Royal Brompton | 1 (4%) |
| Royal Free | 1 (4%) |
| Royal Papworth Hospital | 8 (29%) |
| Royal United Hospital Bath | 2 (7%) |
| Sheffield | 1 (4%) |
| **Research involvement: Yes/No** | 26 (93%)/ 2 (7%) |
| **Patients, relatives, carers N=35** |  |
| **Role:** |  |
| Carer | 1 (3%) |
| Patient | 31 (89%) |
| Relative | 3 (9%) |
| **Centre:** |  |
| Great Ormond Street | 1 (3%) |
| Imperial and Hammersmith | 9 (26%) |
| Newcastle Freeman | 1 (3%) |
| NHS Greater Glasgow and Clyde | 7 (20%) |
| Royal Free | 1 (3%) |
| Royal Papworth Hospital | 5 (14%) |
| Sheffield, Royal Hallamshire Hospital | 11 (31%) |
| **Age at diagnosis** | 45 [34;54] |
| **Age current** | 55 [49;62] |
| **Sex: F/M** | 27 (77%)/8 (23%) |
| **Mutation: Yes/No/UNK** | 12 (34%)/22 (63%)/1 (3%) |
| **N. of research studies in 5 yrs** | 1 [1;3] |
| **Education:** |  |
| Higher degree | 9 (27%) |
| Secondary | 12 (36%) |
| Six Form/College | 12 (36%) |
| **Income:** |  |
| < £12,000 | 6 (18%) |
| £12,000 to £50,000 | 17 (52%) |
| £51,000 to £150,000 | 2 (6%) |
| I prefer not to answer | 8 (24%) |

The Patient and Public Involvement Team of the UNIPHY UK trial network reviewed patient-facing documents for this study. Ethical approval was obtained from the North of Scotland Research Ethics Service (REC: 22/NS/0127). All participants provided written informed consent prior to enrollment in the study. The interviews and focus groups were conducted by MF, a researcher with qualitative research training, who remained blind to the participant’s medical history. Data collection took place between January and August 2023 through telephone or Zoom interviews, lasting between 30 and 60 minutes (Supplement). The interviews were recorded and transcribed verbatim while ensuring anonymity and accuracy. Following the principles of Grounded Theory^9^, the interviews were thematically coded using inductive analysis. The coding process was conducted independently by the first and second authors using MAXQDA (2022), with regular discussions to establish consensus. Partitioning around medoids clustering was performed in R on Jaccard similarity matrices based on the existence of code. The reporting of the study adheres to the COREQ^10^ and SRQR standards^11^. Names, specific locations, and any other identifying details have been omitted to ensure the confidentiality of the individuals involved. Furthermore, the language in certain quotes has been modified to remove explicit references to the patient’s origin or specific cultural background.

In this paper, we focus on the researchers’, clinicians’ and study team’s perspectives.

## Results

### Successes and challenges

The interviews yielded several key themes that shed light on various challenges and successes in genetic research within the context of the NBR and the PAH Cohort study. These themes encompass study design and its evolution, consent procedures and data use, the return of individual genetic results and its impact on clinical practice (Supplement Table 1).

The study can be said to have made substantial contributions to advancing genomic knowledge in pulmonary arterial hypertension (PAH), with clinicians, researchers, and the study team commending its identification of rare and common genetic variants associated with PAH and the discovery of disrupted pathways in the condition. Efficient utilisation of existing NHS infrastructure, standardised healthcare practices, and streamlined logistics were recognised as pivotal in the study’s success. It directly influenced clinical practice by shaping PAH gene panel selection and improving clinicians’ understanding of clinical genetics. The study fostered collaborative networks and interdisciplinary partnerships, a crucial aspect given the rarity of the disease and the small PH scientific community. Challenges included lengthy R&D processes and delays in contract negotiations. Varying clinician engagement levels highlighted the need for a forward-looking approach to research projects. Knowledge gaps and communication barriers underscored the importance of mutual understanding. Data collection challenges involved manual data collection, high missingness rates, and a lack of preparedness for pandemic situations. Delayed return of genetic results and recruitment of relatives posed challenges, impacting family members’ understanding and participation.

### Contribution to science

The NBR and PAH Cohort studies were commended for their substantial contributions to advancing genomic knowledge in PAH, in particular its ability to identify new genes which *“have subsequently been identified as disease-causing. (Clinicians>DS305)* Clinicians, researchers and the study team specifically praised the identification of new rare and common genetic variants associated with PAH, as well as the discovery of disrupted pathways in the condition.

Additionally, the study was prolific in publishing these discoveries and advancements, with one participant saying they have “*lost count of how many papers have been published from the NBR and PAH Cohort with new biomarkers, new ways of diagnosing patients early.” (Study Team>DS327)*

Furthermore, they acknowledged the study’s positive impact on academic training: *“[The study] fostered a whole number of PhD students who rely on the PAH Cohort for their studies” (Study Team>DS327)* and appreciated the wider accessibility of study data through platforms such as the GWAS catalogue and by request for rare variant data.

> *“I just deposited the results of the common variant analysis to an online catalogue. It’s called the GWAS catalogue, and so those data are now freely available for anyone to use for any research that people want to run on the genetics of pulmonary hypertension. So, that’s nice as well because that’s a resource for the community, which hopefully will be useful for lots of different studies when you can use the common genetic associations to test different hypotheses.” (Researchers>DS328)*

### Enhancing existing NHS infrastructure for genetic testing

Clinicians, researchers, and the study team emphasised that the efficient utilisation and enhancement of existing infrastructure were pivotal drivers of the study’s success. Notably, the UK boasts nine Pulmonary Hypertension (PH) centres, which follow standardised protocols. Many of the clinicians have a history of training together, fostering a collaborative professional network. This close-knit community facilitated the smooth integration of the study within the existing healthcare framework, enabling standardised approaches to patient care and research practices across multiple PH centres. This cooperative atmosphere played a significant role in streamlining logistics, data management, and patient recruitment, ultimately contributing to the study’s successes.

Additionally, they acknowledged the centralised nature of the UK’s NHS and its pivotal role in establishing a robust research framework. The support and funding provided by the NHS’s research infrastructure and networks, including the NBR, were highly valued. Participants also commended the study for its non-intrusive nature, seamlessly running alongside their routine clinic visits.

> *“Also, just kind of by the way that the NHS works that everything is connected up and centralised that the pulmonary hypertension service is already set up in a way where all of the centres talk to each other, and they have standards, and they have a history of working together and things, but this study, in particular, helped them to come together because it came with sort of the core facilities of having a place to collect samples, people to organise it, people to chase them, and it provided funding for nurses at the centres to collect the samples.” (Resaerchers>DS328)*

In conclusion, the efficient utilisation of existing infrastructure and collaboration among PH centres were key factors in the study’s success, streamlining patient care, research practices, and logistics.

### Clinical impact and genetic counselling

The study’s results were almost uniformly seen as directly influencing clinical practice. Clinicians and the study team acknowledged the study’s impact on shaping the selection of the PAH gene panel used in the clinical setting, exemplified by initiatives such as PanelApp and on the shape of larger national initiatives such as the NHS Genomic Medicine Service.

> *“Well, for us, it helped us further develop the UK research network, so definitely, that’s grown a lot from NBR and Cohort. […] we got more used to genetic testing, so that’s good. Now, we can do it much more easily on the NHS. So before NBR, we just ignored it all; now we do it routinely. ” (Clinical Teams>DS344)*
>
> *“There are new treatments now that are being explored, the pathways that are being identified on a genetic level. So, I think it’s managing to bring this to another level now at the moment. I’m sure there will be more to come over the next few years.” (Clinical Teams>DS345)*

Interviewees noted that over the course of the study, clinical teams developed a better understanding of clinical genetics and acquired effective communication skills, enabling the integration of genetics into clinical practice.

> *“The results from the sequencing were confirmed and fed back to the patient. So, I think that whole process was developed specifically for the PH patients as part of this, and I’m sure the PH Consultants that you’ve talked to have improved the way that they’re able to communicate these kinds of results with confidence.” (Reserachers>DS328)*

Respondents praised the establishment of pathways that successfully delivered research results to patients, and they appreciated the practical application of the study’s findings through genetic counselling provided by participating geneticists in clinical settings.

> *“So, I went to the genetic clinic for one day and just sat with a counsellor, watching them talk to people who’ve had some genetic testing done, and they were there to get the results and just to see how they dealt with it, how the patients reacted to the news, just things like that really.” (Clinical Tams>DS348)*

Some clinicians remarked that they envision fully integrating genetic testing within their practice as has happened in other diseases, rather than relying only on ad hoc genetic services.

### Fostering collaborative networks

Respondents acknowledged the study’s role in promoting collaborative networks and interdisciplinary partnerships, which significantly contributed to advancing research in the field, with one clinician summarising it as follows, “*the community that is built around it, is probably as valuable, if not more valuable, than the resource itself.” (Clinical Teams>DS317)*

They also emphasised that it is essential to work collaboratively in order to secure the necessary research funding, especially given the rarity of the disease. The study’s facilitation of national and international collaborations, including initiatives like PAH ICON and the UniPHy Clinical Trial Network, was highly valued by both clinicians and researchers.

Overall, respondents identified these successes as key outcomes of the study, highlighting its substantial contributions to science, optimised infrastructure, clinical impact, and the fostering of collaborative networks.

### R&D processes and contractual considerations

Setting up the study presented various challenges for both the study team and some study sites, especially in the realms of R&D processes and contract negotiations. In the R&D phase, the study team encountered resource constraints for the second phase of the PAH Cohort study,

> *“I think I started 20 years ago, and the wiggle room in the service was unbelievable. I dread to think what it’s like now post-pandemic. I mean, to bring a relative in and do all those things. It’s lucrative doing research for hospitals, but I don’t know what it’s like now, whether they have the capacity to do it.” (Study Team>DS339)*

Some researchers mentioned limited funding being a hurdle to relative recruitment and added that *“most things can be solved with more money, right? [If] they’d had more money, every centre could’ve hired someone else who could’ve focused more on relatives, and that could’ve improved things.” (Researchers>DS328)*. The absence of a dedicated data management staff member was also seen as an impediment to efficient data sharing.

Additionally, the process of negotiating contracts with participating institutions proved to be time-consuming, leading to delays in study initiation and, in one instance, resulted in a centre choosing not to participate at all. Similar challenges were faced in negotiating contracts with industry partners, some of which ultimately led to unsuccessful collaborations.

Whilst these challenges did not prevent the successful delivery of the study, they highlight the importance of addressing funding limitations, ensuring adequate resources, and streamlining contract negotiations to facilitate successful study implementation.

#### Variable interest and engagement among participants

Clinicians’ varying clinical and other commitments led to differing levels of interest and engagement in the study; one study team member summarised it as follows, *“changing the culture from a service delivery perspective to one of a research perspective was really challenging throughout the whole study. (Study Team>DS327)*

The lack of actionability of research findings discouraged some clinicians from engaging, highlighting the importance of effectively communicating the clinical relevance of the study. Others highlighted that *“people overestimate what can be done in the short-term, but then they probably underestimate what can be done in the longer term”. (Clinical Teams>DS317)*

and that a forward-looking approach is needed in research projects like this. Interestingly, patients and carers were uniformly not concerned about the limited practical applicability of research findings, highlighting that the findings may help others in the future. One of the study team members summarised this dissonance as follows,

> *“The biggest inertia was from the clinicians rather than the patients. We conducted a study in collaboration with the PHA UK, and there is a questionnaire available on their website that you may have seen. In response to a specific question about whether patients would want to be considered for genetic testing if there was a genetic basis to their disease, around 75-80% answered yes.” (Study Team>DS327)*

### Knowledge gaps, communication barriers, and evolution of the study

Knowledge gaps and varying levels of expertise in research, clinical medicine and genetics impacted study delivery and resulted in communication barriers between stakeholders. Despite all participating centres receiving uniform resources and training in the areas of consent and genetic counselling, variations in local service infrastructure and access to genetic services have surfaced, leading to differences between the centres. These disparities, in turn, have had a broader influence on clinicians’ confidence in delivering genetic results and their overall engagement in the study. To mitigate this, some clinicians have expressed a strong desire for additional training, the establishment of common approaches and resources, as well as the development of standardised patient-facing documents.

Given that the study involved specialists from diverse disciplines, communication barriers have consistently emerged as a root cause of varying levels of engagement. One clinician commented, *“So, you need to make that extra space to learn each other’s language because then I think you also don’t value the contribution of each of the members equally […]. If we get a better understanding of it, we might get better research questions.” (Clinical Teams>DS317)*

Researchers and the study team also highlighted that the study had a limited number of geneticists, leading to a learning process rather than relying on pre-existing genetic expertise.

> *“I guess one of the broader challenges was although we were doing genetic research, probably actually the number of people who were geneticists by background involved in the study were fewer than you might expect, or they had smaller roles than you might expect. […] But it was a challenge in that respect that there wasn’t always somebody who knew exactly what we should do and who was guiding everything. There was a bit of learning on the job, for sure” (Researchers>DS328)”*

Several clinicians, researchers and study team members highlighted that the study had undergone notable evolution over time, with several key developments shaping its trajectory, including shifting from WES to WGS, expanding the number of clinical data points collected, modifying the observation period and visit frequency, introducing remote consenting and saliva along with blood sampling. Over time, the study witnessed a shift from a purely research-focused to a more clinically oriented endeavour. However, this transition has revealed a lack of support and structures initially in place for the clinical aspect (genetic counselling and confirmation of the research findings in the NHS laboratory). Ultimately, there has been a change in the mindset regarding the utility of genetic knowledge,

> *“So, it was a complete mindset change, the PAH clinical space, clinicians, they did not think about these things very much. Unless there was a family sitting in front of them who obviously, two or three family members had the disease, which is the minority of the genetic causes of people with PAH, so that was very challenging.” (Study Team>DS327)*

Finally, all clinicians admitted to initially finding the process of feeding back the results challenging, stating that whilst *“obviously [we] are experts in pulmonary hypertension, [we are not experts] at explaining genetics or the[ir] implications” (Clinicians>DS295)* on various aspects of life, ranging from life insurance to reproduction.

These findings suggest that, overall, the RAPID-PAH study revealed a steep learning curve for both clinicians and researchers, exacerbated by the evolution of the NBR and PAH Cohort studies over time. This underscores the importance of effective communication and knowledge transfer throughout the study’s progression.

### Data collection and sharing

The NBR and PAH Cohort studies aimed to establish a collaborative network to comprehensively investigate the genetic and environmental factors in PAH. Long-term follow-up of patients and relatives would offer insights into genetic mutation effects and treatment responses. This knowledge would lead to more accurate risk estimates for family members and innovative treatment approaches. This involved creating a diverse sample biobank and collecting computable phenotypic and survey data. While this endeavour was largely successful, several challenges were identified by clinicians, researchers and the study team, revolving around the manual collection of non-exclusive data sets (high number of variables), high missingness rates, and absence of electronic consent or non-invasive DNA collection methods from the beginning of the study. A lack of preparedness for pandemic situations exacerbated these issues, *“the second five years, we pretty much got funding to do the same thing, and we didn’t sort of smarten it up a bit in terms of, and I think if we had have been smarter, COVID may not have had such a big impact on us”. (Study Team>DS327)*

However, several members of the study team acknowledged that the pandemic became a catalyst for positive change, paving the way to normalising, for example, the use of digital technologies for online consultations and remote follow-up. One researcher highlighted the lack of efficient data-sharing protocols and indicated that a dedicated team for data cleaning and distribution could be a potential solution and recognised that ultimately, *“data sharing is one of the best ways of doing quality control”. (Researchers>DS328)*

### Delayed or lack of return of individual genetic results and recruitment of relatives

The delayed return of individual genetic results caught many clinicians off guard, leaving them feeling initially unprepared to effectively communicate these results to their patients. *“I suppose some of the frustration was to get some of the genetic results back. It took quite a long time, and you know, in retrospect, that was maybe something that was quite frustrating about that, and then, of course, we have to get the clinical confirmation of that.” (Clinicians>DS349)*

This challenge was even more pronounced in centres without immediate access to genetic services. The clinical teams expressed concerns regarding the delayed return of genetic results, as some patients either had no recollection of enrolling in the study or had died. This complicated the process of delivering genetic results to their family members, thereby affecting their comprehension and ability to make informed decisions.

Clinicians were unaware of the disappointment patients experienced due to the absence of feedback regarding negative genetic results or the lack of updates on the study’s progress.

Recruiting relatives for the study presented several challenges for various reasons. Firstly, contacting relatives could only commence once probands were identified as carrying a deleterious variant. Consequently, the recruitment of relatives was hindered by delays in returning individual genetic results to probands, as pointed out by one participant,

> *“I guess the design was that for relatives to be involved, there was always a lot of uncertainty about how they became eligible to be in the study, and then they only wanted relatives of people who had genes of interest. But if you didn’t get feedback, then we weren’t approaching relatives, so we didn’t do very well on the relative side of things just because we never got any, well we had no one who could distinctly say we have a potential heritable background, there’s no family history. So there was very little reason to approach relatives.” (Clinicians>DS348)*

Secondly, poor integration of otherwise healthy relatives within the NHS system, coupled with their geographical dispersion, presented recruitment difficulties. Additionally, the implementation of new care pathways as a consequence of the study enabled local NHS testing and follow-up for patients, which had a less favourable impact on centralised recruitment efforts. Conversely, some clinicians expressed concerns about service capacity constraints if they were required to accommodate a substantial number of healthy carriers over extended periods.

> *“Are we creating more hassle, more uncertainty for patients’ relatives when we don’t have a definite way of identifying these patients early and certainly not one that’s easy to achieve within our service? If we identify lots of patients’ relatives, would we have the capacity to follow them as perhaps we should do?” (Clinicians>DS344)*

These challenges were not encountered in paediatric centres, where families often attend together, and trio analysis is a common practice. Strong familial bonds between parents and children also facilitated recruitment.

> *“They recruit relatives left, right and centre because obviously you’ve got the child coming in, and you can recruit the parents because they are there, and they want to help. And obviously, you can recruit siblings as well. They want to do as much as they possibly can. ” (Study Team>DS339)*

Patients and their relatives reported that personal factors such as busy schedules, anxiety, low personal stakes in research and patients’ own risk-benefit analysis influenced the lack of engagement with the study.

The COVID-19 pandemic further worsened recruitment in all but paediatric centres,

> *“We were very lucky at GOSH to be allowed to continue our research throughout Covid; other centres were not able to, we were. So that was really good, that really helped with recruitment.” (Clinical teams>DS303)*

Researchers acknowledged the need for better-formulated research questions and tools to understand disease mechanisms in relatives and added that the study’s financial constraints could have contributed to the relatively small number of relatives being recruited to the study.

In summary, the studies not only successfully advanced our understanding of the genetic basis of PAH but also established a lasting collaborative network that extended beyond the UK borders. This network laid the groundwork for various collaborative initiatives, including the Uniphy Clinical Trials Network and PAH ICON. The significance of this multidisciplinary collaborative network is underscored by the expertise it has amassed. From a clinical perspective, these studies played a pivotal role in shaping new services and clinical pathways, integrating genetic diagnostics into routine diagnostic workups.

Despite these successes, participants identified several challenges, such as inefficient R&D and contract processes, variable interest among participants, knowledge gaps, difficulties related to data collection and sharing, and delays in returning individual genetic results and recruiting relatives. As the study addressed numerous questions, it paved the way for new research avenues, which were appreciated by clinicians and researchers alike. To capture this sentiment, one clinician stated, *"I don’t want there to be an after-Cohort. Continuation of, you know, a morphing, but I think it’s short-sighted to think that you can collect biobank samples for five or ten years and then close the study down and have all your questions answered. I think that it needs to continue in some shape or form.“ (Clinical Teams>DS301)*

### Next steps and future directions

When discussing the future directions of genetic research in PAH, strong leadership was identified as essential to guide and coordinate future research efforts, ensuring continuity beyond the current NBR and the PAH Cohort study. There was a consensus that the research should continue to validate and confirm findings obtained through computational analysis and translate ‘omic’ results into clinically relevant biomarkers and potential treatment targets. Additionally, testing blood samples at different time points and expanding research beyond blood to other non-invasively obtainable materials was seen as a source of additional valuable data. Exploring new technologies and extending research questions to other types of PH were seen as other avenues worth attention. Remote data collection methods were also acknowledged as a means to gather information from patients in a more accessible and convenient manner. Ensuring accessibility and sharing of phenotype and genetic data were deemed crucial for advancing research and promoting collaborations.

> *“[we have] to curate the data into a more open and friendly useable system that’s more easily interrogated by other people centres, research fellows, you name it, and somebody across the UK would have to be the sort of custodian of those data. […] I don’t think we need to make it any bigger now. We’ve got so much data, so many samples, and so much longitudinal data that it’s there now as a resource” (Study Team>DS327)*

The stakeholders stressed the importance of sharing expertise and knowledge among researchers and clinicians, enabling more comprehensive and impactful studies. Moreover, fostering national and international collaborations was seen as a way to facilitate knowledge exchange, resource sharing, and a broader understanding of PAH genetics. The potential for academia-industry collaboration was also highlighted: *“Academics can only take things so far, they can only discover so much and, yes, but actually, at the end of the day, you need pharma to come in and actually make it happen.” (Study Team>DS327)* These considerations reflect the perspectives of stakeholders regarding the future development of genetic research in PAH. Collaboration, technological advancements, and data sharing were recognised as essential components for advancing our understanding and treatment of this complex disease.

Based on this in-depth analysis, we drew valuable lessons that can be applied to similar studies in the field, contributing to the body of knowledge on this subject (Table 2).

**Table 2.** Top 10 lessons learned from the NIHR BioResourse Rare Disease study and National Cohort Study of Idiopathic and Heritable PAH.

| 10 top lessons learned |
| --- |
| **1. Utilise existing healthcare infrastructure to maximise efficiency and effectiveness of research efforts.** |
| **2. Establish a collaborative network:** Collaborate with multiple centres and leverage global networks for funding applications, a larger patient pool, increased statistical power, and result validation across diverse populations and streamline patient recruitment to avoid competition. |
| **3. Invest in training and resources:** Provide training and resources to clinical teams, especially in areas where expertise may be lacking, to ensure accurate data collection, interpretation, and dissemination of study results. |
| **4. Build trust and engage with patient advocacy groups:** Trusted clinical teams are the best recruiters. Collaborate with patient advocacy groups to raise awareness, facilitate patient recruitment, ensure patient perspectives are considered and disseminate the study results. |
| **5. Prioritise communication and engagement:** Maintain open and transparent communication with participants, clinical teams, and researchers throughout the study, providing regular updates, reiterating goals and addressing concerns. |
| **6. Consider the impact on participants' lives:** Address participant burdens, mitigate time commitments, and emotional impact. |
| **7. Establish a central sample repository:** Creating a central sample repository facilitates efficient storage and access to biological samples, enabling future research and collaborations. |
| **8. Address data management challenges:** Develop robust systems for data management, including secure storage, standardised data entry procedures, and efficient data sharing protocols to ensure accurate and accessible data for analysis. |
| **9. Embrace technological advancements:** Incorporate innovative technologies such as wearable devices, remote monitoring, and electronic consent to streamline data collection, improve patient engagement, and enhance study efficiency. |
| **10. Learn from study limitations:** Reflect on and address study limitations, such as recruitment difficulties and data management issues, to improve study efficiency. |

## Discussion

Advancements in genetic technology, coupled with the demand for more effective treatments and the increasing interest among patients in genetic and genomic investigations, have driven the integration of genomic technologies into clinical practices across a spectrum of respiratory conditions, ranging from cystic fibrosis to lung cancer. However, within respiratory medicine, the rapid pace of technological evolution has not been uniformly matched by the levels of public and patient awareness nor by the proficiency of healthcare professionals in genomics. Recognising the significance of robust research data in the successful clinical implementation of new medical technologies, our objective was to delve into stakeholders’ perspectives regarding genomic research within the context of PAH and the implementation of results in daily work routines.

Our study highlights the intricate dynamics within genetic research, where the motivations and viewpoints of stakeholders can significantly diverge. It underscores the importance of understanding and addressing these distinct perspectives to foster effective collaboration and engagement among all parties involved. In doing so, the study strengthens and expands upon the insights previously gathered through surveys^12^. Furthermore, this emphasis on collaborative understanding and co-creation plays a vital role in bridging the evidence-practice gap. Co-creation processes involving stakeholders from various backgrounds enable the customisation and contextualisation of genetic research findings. This ensures that the research aligns more closely with the real-world needs and expectations of clinicians, researchers, and patients alike, ultimately facilitating the translation of evidence into practice. Likewise, recognising the role of trust, patients’ disease experience and family dynamics can provide valuable insights in tailoring approaches to recruitment to genetic studies.

Similarly, identifying and addressing modifiable barriers to participation can increase stakeholders’ engagement. Beyond shedding light on stakeholder dynamics, this study also provides valuable insights into the complexities that arise at the intersection of research and clinical care. It sheds light on the challenges that research findings may pose for clinicians and their potential impact on altering clinical practices.

By thoroughly analysing the successes and challenges experienced by stakeholders, we provide a comprehensive compilation of lessons learned and future directions for this study and similar endeavours. Collaboration between centres has emerged as a critical factor in the success of these studies, facilitating the collection of extensive and comprehensive data while fostering the development of a network for future research initiatives such as PAH-ICON and uniPHy Clinical Trial Network. Moreover, it is worth noting that collaborative projects in rare diseases, like PAH, often have a higher likelihood of obtaining funding, given the relatively low number of patients, collective expertise and resources of participating institutions, which further strengthens the network’s capacity for impactful research outcomes. Additionally, by streamlining patient recruitment efforts and avoiding competition among collaborating centres, we ensured a more efficient and coordinated approach. The role of the centralised NHS is critical. The NHS has established research infrastructure and networks, including the Rare Disease Collaborative Networks^13^ and the NIHR BioResource Rare Diseases^14^, which provide vital expertise, support and funding for studies in rare diseases. Standardised NHS practices and protocols ensure consistency across study sites, enabling efficient coordination of logistics, recruitment, and data management. Furthermore, international collaboration is imperative to enhance the sample size, particularly in rare diseases, enabling the combination of expertise and diversification of sample characteristics.

Investing in comprehensive training and resources for clinical teams emerged as a pivotal lesson, ensuring successful recruitment and enabling accurate data collection, interpretation, and dissemination of study results. While support in areas where expertise may be lacking was sought within the study network, rare disease education remains an unmet need^15^. Initiatives such as NHS England Genomics Education, Eurordis - Rare Disease Europe in the EU, or the Rare Disease Clinical Research Network in the US may provide additional resources and expertise.

Effective participant recruitment and retention are significant challenges in any study. Trust emerged as a key factor in driving patient participation. Furthermore, establishing a strong rapport between clinical teams and patients enabled the early identification of challenges that participants in genetic and other research may encounter, allowing for the timely implementation of mitigation strategies. As such, open and transparent communication with participants, clinical teams, and researchers throughout the study, including regular updates, addressing concerns, and fostering engagement, was viewed as crucial for participant retention.

Establishing a central sample repository has been shown to be a crucial component of successful research endeavours. A central repository allows for the efficient storage and accessibility of biological samples, providing researchers with valuable resources for future investigations and collaborations; this has been confirmed across multiple rare disease studies^16^. In addition, addressing data management challenges is essential for ensuring the accuracy and accessibility of research data. Developing robust systems for data management, including secure storage, standardised data entry procedures, and efficient data sharing protocols^17^, is essential for maintaining data integrity and enabling effective analysis. Similarly, embracing technological advancements such as wearable devices, remote monitoring, and electronic consent streamlines data collection, improves patient engagement, and enhances overall study efficiency and has been long viewed as a path forward in rare diseases^18^.

### Strengths and limitations

This is the first comprehensive qualitative study that examines the attitudes towards genetic research in PAH among a high number of stakeholders responsible for the design, conduct and analysis of the genetic study. Through in-depth interviews, it uncovers the perspectives on the successes, challenges and future directions of such studies. Additionally, it formulates valuable lessons learned that can be applied to similar research endeavours as well as inform changes to clinical practice. By employing a substantial and representative sample size for qualitative research, the study ensures that a comprehensive and diverse range of perspectives and attitudes are captured, resulting in a well-rounded overview.

Notwithstanding these strengths, the study inevitably has limitations. Firstly, while the study captures a diverse range of perspectives on genetic research, the findings may not be applicable to all genetic research contexts, i.e. common diseases. Secondly, the study focuses on insights from studies performed in the UK, which were deeply embedded in the NHS and may not be fully applicable in other healthcare models or under different regulatory circumstances.

## Conclusion

This analysis has shed light on the importance of collaborative efforts and reflective training within the study; it identified key lessons learned and highlighted promising directions for future genetic research in PAH. By continuing to explore the genetic factors contributing to PAH, we can pave the way for improved diagnosis, treatment, and, ultimately, better outcomes for individuals affected by this complex condition. It is worth noting that the NBR and the PAH Cohort studies stand as unique initiatives in the field of pulmonary medicine, serving as potential transferable examples for research of other rare diseases to follow.

## Data Availability

Anonymised and analysed data from this study can be made available for collaborative work upon request. However, individual data cannot be provided.

## Acknowledgements

We extend our gratitude to the Patient and Public Involvement Team of the UniPHy Clinical Trial Network for their invaluable assistance in reviewing patient-facing documents for this study. We thank all the patients, their families, clinical and research teams who contributed to this research and the Pulmonary Hypertension Association (United Kingdom) for their support. We thank all the participants, the research nurses and coordinators at the specialist pulmonary hypertension centres involved in the National Institute for Health Research (NIHR) BioResource and the Cohort study of idiopathic and heritable PAH.

## Authorial contribution

EMS conceived the project, analysed the data, and wrote the manuscript. MF conducted the interviews and focus groups, analysed the data and co-wrote the manuscript. NWM supervised the project.

## Supplemental Material

## Methods

### Recruitment

A purposive sample of participants from the NBR and PAH Cohort study was selected with a view to include/represent diverse age groups, genders, and individuals with PAH risk gene mutations. The recruitment process covered all PH centres in the UK and took four distinct approaches:

1. **Local Clinical/Research Team Contact:** Participants were contacted via local clinical or research teams, recruiting individuals from 7 out of 9 centres. The local teams employed two approaches: contacting potential participants directly during clinic visits or through phone and postal communication, with direct contact being the more effective method for participant recruitment.
2. **Referrals from Current Participants:** The recruitment pool was expanded through referrals from current participants. In particular, this approach facilitated the recruitment of relatives of existing participants.
3. **Online Recruitment:** An online recruitment strategy involved calls for participation via email, social media platforms, and the Pulmonary Hypertension Association (PHA) webpage. This method led to the recruitment of patients from 2 centres.
4. **Direct Recruitment of Team Members:** Clinical, research, and study team members were directly recruited from all 9 UK centres.

Active recruitment continued for each centre until the target number of participants had been attained.

### Interview topic guides

ES and MF collaboratively developed detailed, semi-structured interview topic guides. Semi-structured topic guides are carefully designed to keep the interviews consistent while also allowing participants to share their unique perspectives. As such, they played a pivotal role in steering the conversation towards the research objectives, ensuring that critical information was collected. Furthermore, interview guides evolved throughout the research process, undergoing refinement based on feedback from earlier interviews or emerging findings. This iterative approach enhanced the depth and relevance of subsequent discussions, ultimately contributing to a richer and more nuanced understanding of the research subject.

## Results

**Supplement Table 1.** Themes and subthemes. Segment refers to the fragment of text that is isolated for analysis. The themes and subthemes enclosed by a thick border in the table were discussed in this article.

| THEMES AND SUBTHEMES | SEGMENTS | PERCENTAGE |
| --- | --- | --- |
| **Research participation - Enablers** | **400** | **100** |
| Altruism/For the greater Good/Future | 121 | 30.25 |
| Relationship with clinical teams/trust | 99 | 24.75 |
| Process and Communication - Do-ability | 65 | 16.25 |
| Diagnostic journey | 35 | 8.75 |
| Seeking answers | 24 | 6 |
| Self-benefit | 25 | 6.25 |
| Rare disease as a motivator | 14 | 3.5 |
| Knowledge and background as enabler | 9 | 2.25 |
| Improving diagnosis | 2 | 0.5 |
| Community of participants | 6 | 1.5 |
| **Research participation - Barriers** | **139** | **100** |
| Impact on participants' lives | 33 | 23.74 |
| Invasive procedures | 60 | 43.17 |
| Clinical trials: reluctant to rock the boat | 15 | 10.79 |
| Feeling overwhelmed and disempowered | 16 | 11.51 |
| Lack of trust or study done by non respected institution | 5 | 3.6 |
| Insurance | 10 | 7.19 |
| **Study setup** | **613** | **100** |
| Scope, research questions and study design | 33 | 5.38 |
| Recruitment of participants: patients and relatives | 55 | 8.97 |
| Consent | 258 | 42.09 |
| Data and samples (collection/use/storage/process) | 102 | 16.64 |
| Return of genetic results | 117 | 19.09 |
| Resources/Support/Funding | 48 | 7.83 |
| **Study successes** | **178** | **100** |
| Scientific and study community | 62 | 34.83 |
| Outputs and innovation | 78 | 43.82 |
| Clinical impact and infrastructural changes, new care pathways | 27 | 15.17 |
| Expanding knowledge and skills | 11 | 6.18 |
| **Study challenges** | **207** | **100** |
| R&D processes and contractual considerations | 6 | 2.9 |
| Variable interest and engagement among participants | 7 | 3.38 |
| Genetics knowledge/confidence/attitude | 41 | 19.81 |
| Data/Sample collection and sharing | 19 | 9.18 |
| Delayed or lack of return of individual genetic results | 57 | 27.54 |
| Recruitment of relatives | 16 | 7.73 |
| Lack of care pathways or guidelines | 41 | 19.81 |
| Impact of COVID-19 (care/health/recruitment) | 20 | 9.66 |
| **Next steps** | **51** | **100** |
| Stakeholders engagement and training | 8 | 15.69 |
| Data accessibility, collaboration, industry partnership | 9 | 17.65 |
| Exploring new technologies and techniques | 13 | 25.49 |
| Expanding research to other PH groups and sample types | 9 | 17.65 |
| Validating study findings and making them actionable | 4 | 7.84 |
| Leadership, vision, funding | 8 | 15.69 |

